# Ribosomal, Satellite III (1q12) and Telomere Repeat Copy Number Variations in Cystic Fibrosis Patients Used as a Model of Permanent Stress and Survivability

**DOI:** 10.1101/2024.04.21.24306126

**Authors:** E.S. Ershova, E.I. Kondratyeva, L.N. Porokhovnik, A.Y. Voronkova, Yu. L. Melyanovskaya, S.A. Krasovsky, R.V. Veiko, E.K. Zhekaite, M.A. Starinova, N.N. Veiko, S.V. Kostyuk

## Abstract

**Introduction:** Ribosomal (rDNA), satellite III (f-SatIII) and telomere (TR) tandem repeats perform a variety of functions in the human cell. Copy number variations (CNVs) of these repeats contributes to the global chromatin architecture and genome expression changes in response to stress or pathology. An elevated rDNA abundance and lowered contents of f-SatIII and TR were found in blood leukocytes of patients with schizophrenia. The main question of the study was: are the CNVs of the three repeat types in blood leukocytes a universal phenomenon linked to a patho4logy associated with chronic oxidative stress? Cystic fibrosis (CF), a monogenic disease was chosen as an object of the study.

**Materials and Methods:** We determined the rDNA, f-SatIII and TR content in the blood leukocyte genomes of 545 subjects aged 0.2 to 50 years. The subjects were divided into three groups: Control (HC-group, N = 267), CF group (N= 186) and CF(d) group (severe patients, who died after some time upon sampling, aged 17 to 40 years, N = 92). For each patient, the type of the mutation in *CFTR* gene had been determined earlier. Non-radioactive quantitative hybridization technique was applied to quantify the repeats.

**Results:** the rDNA abundance was elevated in the DNA of CF and CF(d) groups (565±105 copies per genome, N=278) compared to HC group (445±112 copies, N=267). A patient’s age 3 to 16 years was associated with “severe” mutations in *CFTR* (98% of cases), increased f-SatIII repeat counts and decreased telomere repeat (TR) contents in genome DNA compared to the age-matched controls. The genomes of deceased patients from CF(d) group also harbored increased numbers of f-SatIII and decreased numbers of TR. Patients above 16 years with a milder course of the disease and relatively low content of “severe” *CFTR* mutations contained less f-SatIII and more TR in their genomic DNA. A parameter **rDNA·(f-SatIII/TR)** showed a maximum difference between patients with relatively mild (age 17 to 40) and severe (age 17 to 40) forms of the pathology according to ROC analysis data (AUC = 0.86).

**Conclusion:** Cystic fibrosis was associated with an increase in rDNA abundance and altered f-SatIII and TR contents in the DNA of cases compared to the controls. The severe course of the disease was characterized with high f-SatIII contents and shortened telomeres. Whereas mild CF cases were associated with low contents of f-SatIII and normal or slightly reduced telomere length. The index **rDNA· (f-SatIII/TR)** might be a predictor of the patient’s life expectancy.

## 1. INTRODUCTION

The human genome contains a significant number of repetitive sequences. Some of the repeats are clustered into tandem arrays. The tandem repeats play a principle regulation and structuration role in the functioning of the nuclear chromatin. Copy number variants (CNVs) of the tandem repeats have been implicated in many diseases. The tandem repeats demonstrate quantitative polymorphism, diversity in their nucleotide structure, localization on the chromosomes, their functions and stability in the processes of aging and stress response [1–11].

The ribosomal genes (rDNA) are a special type of tandem repeats. Human genome contains 100 to 900 rDNA copies per diploid cell [12–15]. A 44838 nucleotide pair long repeated unit contains a transcribed region, which codes for 47S rRNA, and a non-transcribed intergenic spacer, that includes non-extended repeats – microsatellites and transposons [15, 16]. Processing of 47S rRNA provides RNA synthesis (18S, 5.8S, 28S rRNA). The ribosomal tandem repeats are localized in p-arms of the five acrocentric chromosomes [17]. During interphase, the ribosomal repeats form a special structure termed nucleolus (Fig.1).

**Figure 1.**
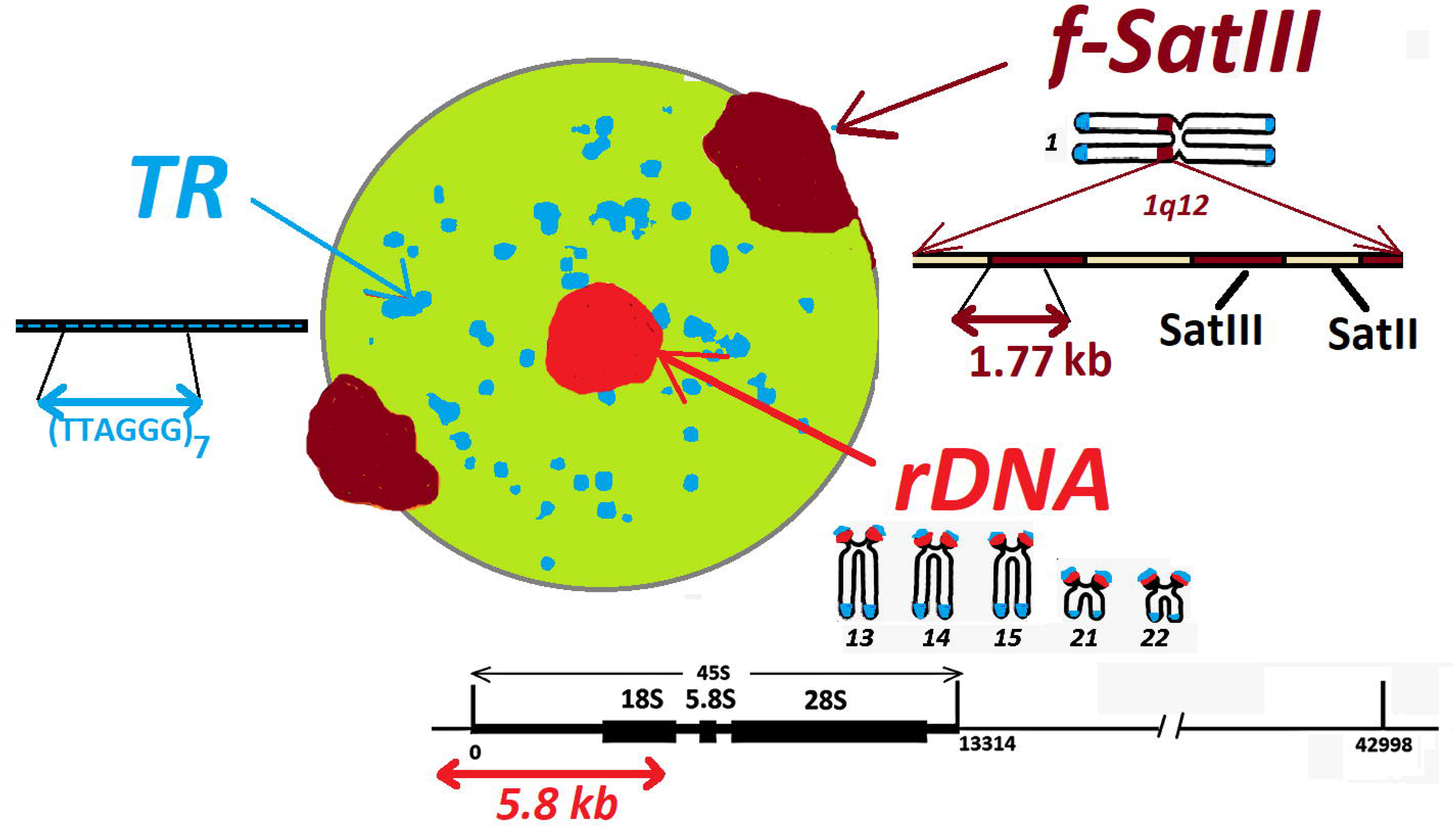
The layout of human genome’s tandem repeats studied. The localization of the repeats in the interphase nucleus is indicated on the base of previous studies [18, 19]. The DNA probes used for hybridization are shown.

The functions of the nucleoli are not limited only to the production of ribosomal subunits [20–22]. The nucleolus is a complex hub, where the processes of ribosome biosynthesis, the cell cycle progress and the response to various types of stresses and viral infections are coordinated. Recent studies have shown that epigenetic status of the ribosomal genes and integrity of the nucleolar structure can modulate the cell homeostasis [23–28]. The discovery of structural and functional links between the nucleolus and the rest of the cell genome has shown that the nucleolus plays a key role in the organization of the nucleus architecture [29]. Copy number variations of the ribosomal repeats change the cell response to DNA damage [30].

A large amount of rDNA in the nucleolus stabilizes the nuclear heterochromatin by blocking the heterochromatin to euchromatin transition [31, 32]. The heterochromatin includes satellite repeats, or satellites. The satellites are tandemly repeated sequences organized into extended arrays and located in the centromeric and pericentromeric regions of the chromosomes. The number of the satellite DNA arrays can vary several times in individual chromatin regions [33]. The satellites play a structural and regulatory role in organizing the chromatin in the nucleus. The critical role of the pericentromeric repeats in maintaining chromosomal integrity during aging has been established [9, 34].

Previously, we studied a fragment of the pericentromeric satellites III (f-SatIII) using the DNA probe PUC1.77 [35] specifically hybridizing to the 1q12 pericentromeric heterochromatin of the first chromosome (Fig.1). A distinctive feature of the f-SatIII is the instability of its content in the DNA of the cell pool under conditions of oxidative stress and aging. Replicative senescence is accompanied by the accumulation of f-SatIII in a fraction of the cells [36]. The cells with high counts of the repeat are unable to divide and do not respond to stress by developing an adaptive response. These cells die when the stress levels rise [18, 37–40]. Transcription of satellite is one of the eventual mechanisms leading to the amplification of this repeat in some cells of a senescent population [41, 42].

The amplification of f-SatIII in senescent cells negatively correlates with a content of the tandem repeat (TTAGGG)n, which forms the telomeres [18, 43]. The telomere regions protect the chromosome ends against agglutination or base pair loss and maintain the stability of the cell cycles [44]. Unlike euchromatin sites of the genomic DNA, repair of single breaks is poorly developed in the telomeres’ DNA [45]. The two major signs of human physiologic aging, namely, loss of genome integrity and telomere shortening, result in disruption of the core architecture and replicative senescence [46]. The telomere shortening is associated with decondensation and transcription of heterochromatin sites [47–52].

We found early an interesting fact: DNA derived from schizophrenia cases contained low copy numbers of f-SatIII repeats together with an increased copy numbers of rDNA (rDNA CN) compared to the controls [43]. A decrease in the content of telomere repeats in patients with schizophrenia was also reported [53, 54]. Schizophrenia is a multifactorial disease that is accompanied by chronic oxidative stress [55]. We believe that oxidative stress is the cause of the observed CNVs of the tandem repeats in schizophrenia patients compared to healthy controls [43].

In order to understand how universal the profile of CNVs of the three repeats is under conditions of chronic oxidative stress induced by the pathology, we analyzed DNA samples isolated from blood leukocytes of patients with cystic fibrosis. Cystic fibrosis (CF) is an autosomal recessive disorder due to mutations in transmembrane conductance regulator (*CFTR*) gene leading to the abnormality of chloride channels in mucus and sweat producing cells. *CFTR* dysfunction causes the dysregulation of several signaling pathways to generate an innate oxidative state. An essential deficit of antioxidant molecules and an increase in oxidative stress have been shown in the patients with CF. The sustained imbalance between oxidant and antioxidant species induces chronic inflammation. The role of oxidative stress in the progression of lung injury in the CF patients has been described [56, 57].

Thus, a common feature of the two fundamentally different diseases (schizophrenia and cystic fibrosis) is systemic chronic oxidative stress, which could cause CNVs of the three tandem repeats in the genomes of blood leukocytes derived from patients compared to healthy controls. In order to test this assumption, we quantified the three repeats in DNA samples isolated from blood leukocytes of 267 healthy controls and 278 subjects with cystic fibrosis.

## 2. MATERIALS AND METHODS

### 2.1. Subjects

Totally, 278 cystic fibrosis cases aged 0.21 to 49 years were studied. Male to female ratio of the total sample was 1:0.96. The cases were divided into two groups. CF(d) group included 92 patients aged 16.8 up to 38.4 years, with a mean of 24.5±5.0 years, who died within several months after blood samples had been taken. CF group included 186 survived cases aged 0.21 up to 49.6 years, with a mean of 17.4±13.5 years.

CF was diagnosed according to the criteria of the CF clinical care recommendation and the CF national consensus guidelines (2019) [58]. For the assessment of patient’s status and description of the clinical picture, the Russian Federal CF Patient Register data were used. The Register data were collected from anamnesis and outpatient records of patients from Russian CF centers. The Register format corresponded to the European Register of CF patients [59]. The study design and informed consent form were approved by Ethics Committee of Federal State Budgetary Scientific Institution Research Centre for Medical Genetics named after academician N.P.Bochkov (RCMG). Each patient was under dispensary observation at the Research and Clinical Cystic Fibrosis Department of RCMG and was treated at the Cystic Fibrosis Departments of the Moscow Region Research Clinical Institute of Childhood and/or Pulmonology Scientific Research Institute under FMBA (Federal Medical and Biological Agency of the Russian Federation).

In the CF group patients, the following statistically significant correlations with the patient’s age were registered: (1) increasing age of diagnosis; (2) increasing frequency of chronic and intermittent infections caused by Pseudomonas aeruginosa, chronic infections by Burkholderia cepacia, Staphylococcus aureus, Achromobacter spp.; (3) decreasing respiratory function indices such as forced expiratory volume in 1 second (FEV1) and forced vital capacity (FVC). Complications such as allergic bronchopulmonary aspergillosis, diabetes mellitus, pneumothorax, hemoptysis, upper respiratory tract polyposis and osteoporosis prevailed in the groups of adult patients, especially among the deceased, compared to groups of children.

The microbiological status of the bronchopulmonary system was assessed using an algorithm for microbiological diagnosis of chronic lung infection in patients with CF, including application of bacteriological, biochemical, and other phenotypic tests (detection of hemolysis, test for biofilm formation capability) and molecular biology methods [58]. The biomaterials used in the medical inspection of the lower respiratory tract in patients with CF were sputum when coughing, a smear from the throat after coughing, laryngeal or nasopharyngeal aspirate, sputum induced by a hypertensive solution, bronchoalveolar lavage, brush biopsy material during bronchoscopy.

Testing for genetic variants of *CFTR* gene was conducted according to the standard algorithm [58, 60]. Most ‘mild’ [60] genotypes (34%) were found in the groups of adult patients over 17 compared to the groups of children (2.1%). In CF(d) group, the fraction of patients with ‘mild’ genotype (32%) was significantly (p <0.001) less than in the age-matched CF group (59%).

As healthy controls (HC group), DNA samples were obtained from the collection of the Laboratory of Molecular Biology of RCMG named after N.Bochkov. The dataset included 267 unrelated individuals (51% male) inhabiting Moscow city and the Moscow region. The HC group included healthy persons aged 5 to 50 with a mean age of 22.3 ± 11.5 years, who carried no mutation in *CFTR* gene and no other diagnosed mutation linked to genetic pathology.

The contents of f-SatIII and TR in human DNA are known to depend on the subject’s age and stress status [37, 40, 43]. In view of this, all patients were divided into five groups:

1. CF(0-2): N=37, the age was from 0.21 to 2 years, the ‘severe’ genotype *CFTR* was detected in 97% of patients from this group;
2. CF (3-16): N=54, the age from 3 to 16 years, the ‘severe’ genotype was detected in 98% of the patients;
3. CF(d): N=92, the age from 17 to 40 years, the ‘severe’ genotype detected in 68% of the patients. All the cases included in the group died within several months after blood sampling, thus indicating a very severe course of the disease;
4. CF(17-40): N=83, the age matched the age of patients from CF(d) group, the ‘severe’ genotype was detected in 58%;
5. CF (41-50): N=12, age from 41 to 50, the ‘severe’ genotype detected in 40%.

The control cohort was divided into three groups, that matched the patient groups by age: HC (3-16), HC (17-40) and HC (41-50).

### 2.2. DNA Samples

5 mL of blood was collected from the peripheral vein using a syringe flushed with heparin (0.1 mL/5 mL blood) under strict aseptic conditions. Leukocytes were isolated from the blood after erythrocyte lysis. The DNA isolation method was described in detail previously (Chestkov et al. 2018). Briefly, 5 mL of the solution (2% sodium lauryl sarcosylate, 0.04 M EDTA), and 150 μg/mL RNAse A (Sigma, United States) were added to 0.5 mL of the leukocyte suspension (37°C, 45 min). The lysate samples were treated with proteinase K (200 μg /mL, Promega, United States) for 24 h (37°C). The samples were extracted with 5 mL of freshly distilled phenol (stabilized with 8-hydroxyquinoline), 5 mL of phenol/chloroform/isoamyl alcohol (25:24:1), and 5 mL chloroform/isoamyl alcohol (24:1). DNA was precipitated (0.3M sodium acetate, pH 5.2, and 2.5 volume of ethanol, −20°C) and collected by centrifugation (10,000 g, 15 min, 4°C), washed with 70% ethanol, then dried, and dissolved in water. The DNA concentration and purity were determined spectrophotometrically. The final DNA quantification was performed using PicoGreen dsDNA quantification reagent (Molecular Probes, Carlsbad, CA, United States). The DNA concentration in the samples was calculated according to a DNA standard curve. EnSpire equipment (Finland) with excitation and emission wavelengths of 488 and 528 nm was used.

### 2.3. Non-radioactive Quantitative Hybridization (NQH)

The method was described in detail in a previous publication [61]. Briefly, 2µL (20 ng/µL) of the denatured DNA samples (4-6 dots/sample) were applied to a filter (Optitran BA-S85, GE healthcare). 2µL of six standard samples of the genomic DNA (20 ng/µL) with a known content of the rDNA, f-SatIII and telomere repeats (TR) were applied to the same filter, in order to plot a calibration curve for the dependence of the signal intensity on the contents of the repeats in a particular sample. The filter was heated (80°C in vacuum, 1.5 h). After hybridization had been completed, the membrane filter was treated with a conjugate of streptavidin-alkaline phosphatase (Sigma) and substrates for alkaline phosphatase (NBT + BCIP). The filter was washed, dried, and scanned. In order to measure the hybridization signal, special software was used (“Imager 6,” RCMG, Moscow). The software determined the dot location and calculated the integral dot intensity. Signals from several dots corresponding to the same sample were averaged. The mean and standard error were calculated. The rDNA, f-SatIII and TR contents in the sample were calculated using a calibration curve equation. Relative standard error for NQH was merely 5±2%. Total relative standard error, that includes standard error for DNA extraction, concentration determination and NQH method, was 11±8%.

### 2.4. The DNA-Probes

Human rDNA pBR322-rDNA probe contains rDNA sequences (5836 bp) cloned into EcoRI site of pBR322 vector. The cloned rDNA fragment covers the positions from −515 to 5321 of the human rDNA (GenBank accession no. U13369), Fig. 1.

F-SatIII probe was a 1.77-kb cloned EcoRI fragment of human satellite DNA [35]. Dr. H. Cook (MRC, Edinburgh, UK) kindly supplied the human chromosome lql2-specific repetitive satellite DNA probe pUC1.77.

The DNA-probes pBR322-rDNA and pUC1.77 were biotinylated with biotin-11-dUTP using a nick translation kit (Biotin NT Labeling Kit, Jena Bioscience GmbH).

For the detection of the human telomere repeat, the probe biotin-(TTAGGG)7 was used. Syntol (Moscow, Russia) performed the synthesis and biotin labeling of the oligo-probe.

### 2.5. Statistical Analysis

From 4 to 6 parallel dots for one DNA sample were applied in one experiment. All reported results were reproduced at least two times as independent biological replicates. The descriptive statistics for rDNA, f-SatIII and TR are presented in Table 1. The significance of the observed differences was analyzed using non-parametric Mann-Whitney U tests and Kolmogorov– Smirnov statistics. Data were analyzed with StatPlus2007 Professional software (http://www. analystsoft.com/). All p-values were two-sided and considered statistically significant at p < 0.001. Descriptive statistics are also provided by Box Plots, which show the median, lower and upper quartiles, the minimum and maximum values, and outliers. MedCalc (https://www.medcalc.org/manual/roc-curves.php) created a complete sensitivity/specificity report using Receiver Operating Characteristic (ROC) curve analysis. Each point on the ROC curve represents a sensitivity/specificity pair corresponding to a particular decision threshold. The area under the ROC curve (AUC) reflects the parameter difference between two groups (disease/health or health/disease).

**Table 1.** Descriptive statistics for the rDNA, f-SatIII and TR content in the DNA of cystic fibrosis patients (CF) and healthy controls (HC).

| DNA | Group | N | Mean | SD | Range | Median | C.V. |
| --- | --- | --- | --- | --- | --- | --- | --- |
| **rDNA**<br>**CN** | CF | 186 | 559 | 118 | 351 - 932 | 538 | 0.21 |
|  | CF(d) | 92 | 565 | 73 | 350 - 892 | 556 | 0.13 |
|  | CF(all) | 278 | 561 | 105 | 350 - 932 | 551 | 0.19 |
|  | HC | 267 | 446 | 112 | 200 - 796 | 442 | 0.25 |
| **f-SatIII**<br>**pg/ng**<br>**DNA** | CF | 186 | 14.6 | 3.8 | 6.4 – 29.0 | 14.0 | 0.26 |
|  | CF(d) | 92 | 20.7 | 7.5 | 8.3 – 50.3 | 19.1 | 0.36 |
|  | CF(all) | 278 | 16.6 | 6.0 | 6.4 – 50.3 | 14.9 | 0.36 |
|  | HC | 267 | 18.2 | 5.9 | 6.0 – 40.0 | 17.0 | 0.32 |
| **TR**<br>**pg/μg**<br>**DNA** | CF | 186 | 310 | 76 | 131 - 535 | 321 | 0.24 |
|  | CF(d) | 92 | 286 | 59 | 110 - 482 | 285 | 0.21 |
|  | CF(all) | 278 | 302 | 71 | 110 - 525 | 300 | 0.24 |
|  | HC | 267 | 356 | 51 | 234 - 544 | 361 | 0.14 |

## 3. RESULTS

### 3.1. Ribosomal repeat CNVs

The cystic fibrosis affected cases differed from the controls by higher values of rDNA CN (Fig. 2Aa1 and Table 1). The patient sample contained no genome with rDNA CN < 350, while 18% of the controls counted less than 350 rDNA copies. The ROC-analysis data (Fig. 2Aa2) corroborated the fact of a significantly higher rDNA content in the genomes of patients with CF. The greatest differences from the controls (AUC=0.87) were observed for the group of deceased patients CF(d).

**Figure 2.**
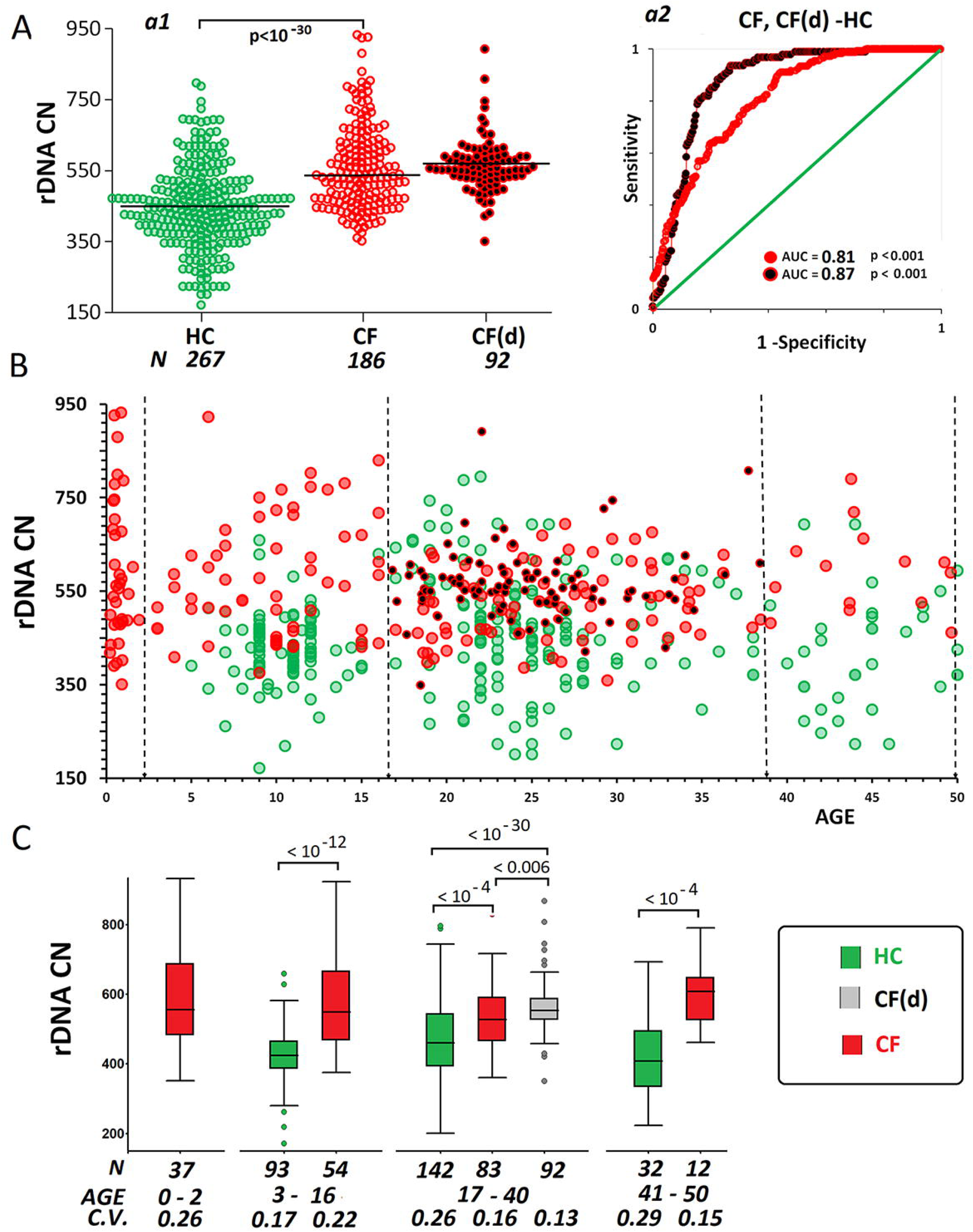
Ribosomal DNA (rDNA) abundance in DNA of cystic fibrosis cases and healthy controls. A. (a1) The rDNA copy numbers in DNA samples from HC, CF and CF(d) groups. A comparison between the groups is reported (U-test). (a2) ROC curves for the groups. The area under the ROC curve (AUC) is a measure of how well a parameter can distinguish between two groups. B. The dependence of rDNA CN on subject’s age in HC, CF and CF(d) groups. The vertical dotted lines indicate dividing into four age groups. C. The rDNA CN analysis in different age groups. Box Plot diagrams and group comparisons (U-test) are shown.

The rDNA CN did not depend on the age in the three groups (p>0.1, Fig.2B and Table 2). In each age group, rDNA CN in patient’s DNA were significantly higher than in the controls (Fig. 2C). The maximum coefficients of variation (C.V. = 0.26 – 0.22) and maximum values of rDNA CN (more than 900 copies) were found in groups of affected children and adolescents. In CF (17-40) and CF(d) groups, reduced intervals and coefficients of variation of rDNA CN were found (C.V.= 0.13 – 0.16). No observation of an rDNA content less than 450 and more than 800 copies occurred in patients over 30 years of age.

**Table 2.** Spearman rank correlations for experimentally determined and calculated parameters.

|  |  | *rDNA* (R) | *f-SatIII* (S) | *TR* ( T ) | *S/T* | *R\*S/T* |
| --- | --- | --- | --- | --- | --- | --- |
| **HC**<br>**N = 267** | **Age** | 0.02<br>0.74 | **0.49**<br>**0.0000** | **-0.32**<br>**0.0000** | **0.55**<br>**0.0000** | **0.39**<br>**0.0000** |
|  | **rDNA (R)** |  | **0.19**<br>**0.002** | 0.07<br>0.25 | 0.15<br>0.010 | **0.64**<br>**0.0000** |
|  | **f-SatIII (S)** |  |  | -0.03<br>0.66 | **0.92**<br>**0.0000** | **0.80**<br>**0.0000** |
|  | **TR (T)** |  |  |  | **-0.36**<br>**0.0000** | **-0.22**<br>**0.0002** |
|  | **S/T** |  |  |  |  | **0.83**<br>**0.0000** |
| **CF**<br>**N = 186** | **Age** | -0.03<br>0.65 | **0.24**<br>**0.001** | 0.02<br>0.74 | 0.13<br>0.09 | 0.06<br>0.38 |
|  | **rDNA (R)** |  | 0.08<br>0.28 | 0.16<br>0.03 | -0.07<br>0.36 | **0.46**<br>**0.0000** |
|  | **f-SatIII (S)** |  |  | 0.01<br>0.85 | **0.65**<br>**0.0000** | **0.60**<br>**0.0000** |
|  | **TR (T)** |  |  |  | **-0.68**<br>**0.0000** | **-0.52**<br>**0.0000** |
|  | **S/T** |  |  |  |  | **0.81**<br>**0.0000** |
| **CF(d)**<br>**N=92** | **Age** | -0.05<br>0.62 | 0.06<br>0.59 | 0.02<br>0.84 | 0.05<br>0.66 | 0.06<br>0.57 |
|  | **rDNA (R)** |  | 0.14<br>0.19 | **0.40**<br>**0.0001** | -0.07<br>0.48 | 0.19<br>0.08 |
|  | *f-SatIII*<br>(S) |  |  | 0.31<br>0.003 | 0.83<br>0.0000 | 0.83<br>0.0000 |
|  | *TR*<br>(T) |  |  |  | -0.18<br>0.09 | -0.09<br>0.38 |
|  | *S/T* |  |  |  |  | 0.95<br>0.0000 |

### 3.2. CNVs of f-SatIII repeats

Fig.3Aa1 and Table 1 represent data on f-SatIII repeat abundance in DNA isolated from blood leukocytes of all subjects studied. The f-SatIII contents in DNA samples that belong to HC-group are significantly higher than those in DNA of patients from CF-group. In CF(d) group, the repeat copy numbers in DNA are more than in HC-group (p < 0.005) and significantly exceed the values in CF group. The differences are substantiated by ROC analysis data (Fig.3Aa2).

Fig.3B shows the dependence of f-SatIII content in DNA of the study participants on their age. The f-SatIII contents of HC-group positively correlated with the subject’s age (Table 2, Rs = +0.49, p<10^-7^, N = 267). The leukocyte DNA of healthy children and adolescents under 16 contained low amounts of f-SatIII (14 ± 2 pg/ng DNA, N=93) compared to subjects at the age of 17 to 50 years (21 ± 6 pg/ng DNA, N=174). The contents of f-SatIII in total DNA samples of the whole CF cohort correlated with the patient’s age to a lesser extent (Table 2, Rs = +0.24, p<10^-3^, N = 186). In the subgroup of affected children 3 to 16 years of age, 23% of DNA samples carried more repeats, than any sample of the age-matched group of healthy children (Fig.3B).

**Figure 3.**
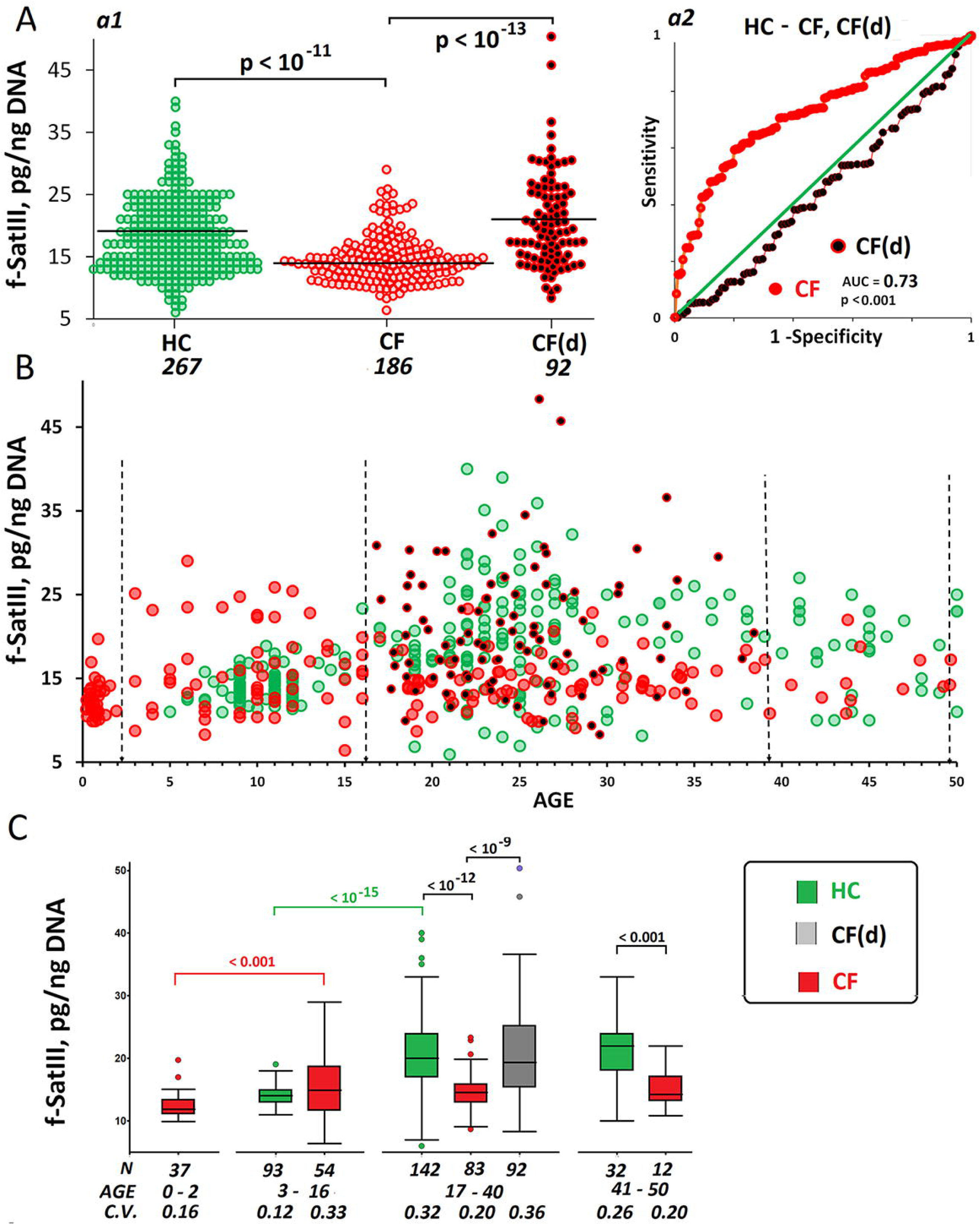
Satellite III (f-SatIII) abundance in DNA of cystic fibrosis cases and healthy controls. A. (a1) The f-SatIII copy numbers in DNA samples from HC, CF and CF(d) groups. A comparison between the groups is reported (U-test). (a2) ROC curves for the groups. B. The dependence of f-SatIII repeat copy numbers on subject’s age in HC, CF and CF(d) groups. The vertical dotted lines indicate dividing into four age groups. C. The f-SatIII CN analysis in different age groups. Box Plot diagrams and group comparisons (U-test) are shown.

In figure 3C, one can see the data of f-SatIII quantification in DNA of the three groups (HC, CF and CF(d)) within the different age intervals. The maximum differences in the repeat content between patients with CF and healthy controls were found in age intervals over 16 years. In CF(17-40) and CF(41-50) groups of these age intervals, the coefficients of variation of the parameter were significantly reduced compared to the subgroups CF(d) and HC.

### 3.3. Telomere repeat CNVs

Fig.4Aa1 and Table 1 represent TR content data for HC, CF and CF(d) groups. The CF cohort differed from the CF(d) subgroup by a higher TR content. The TR content in DNA of cases was considerably lower than in the controls. The differences are corroborated by ROC-analysis results (Fig.4Aa2).

Fig.4B and Fig.4C show a dependence of TR content in DNA on the subject’s age. In HC group, a negative correlation of TR content in DNA with age was observed (Table 2, Rs = −0.32, p<0.0001, N = 267). The TR content in DNA of cases depended non-linearly on the patient’s age and could be described by an equation of the second degree (y = 0.18x^2^ – 6.8x + 346, R² = 0.2). The contents of the telomere repeats were maximum in the infant CF(0-2) group and considerably decreased in CF(3-16) group. An increase in TR content was observed in patients over 16 years of age. In patients who survived to an age of 40+, the TR content did not differ from the repeat content in the controls. The TR content in DNA samples of CF(d) group was lower than in DNA of age-matched CF cases (Fig.4C).

### 3.4. Comparison of CNVs of the three repeat types

Fig. 5 (A-C) represents the results of pairwise comparison of the contents of the studied repeats in cases and healthy controls. Table 2 shows the correlation dependencies. Fig.5D displays a three-dimensional plot showing the contents of the three repeats in DNA samples. In the control group, a weak positive correlation was found between the contents of rDNA and f-SatIII (Table 2, Rs =+0.19, p=0.002, N = 267). In the group of died patients with severe disease course CF(d), a positive correlation was found between the contents of rDNA and TR (Table 2, Rs =+0.43, p<0.0001, N = 92).

**Figure 4.**
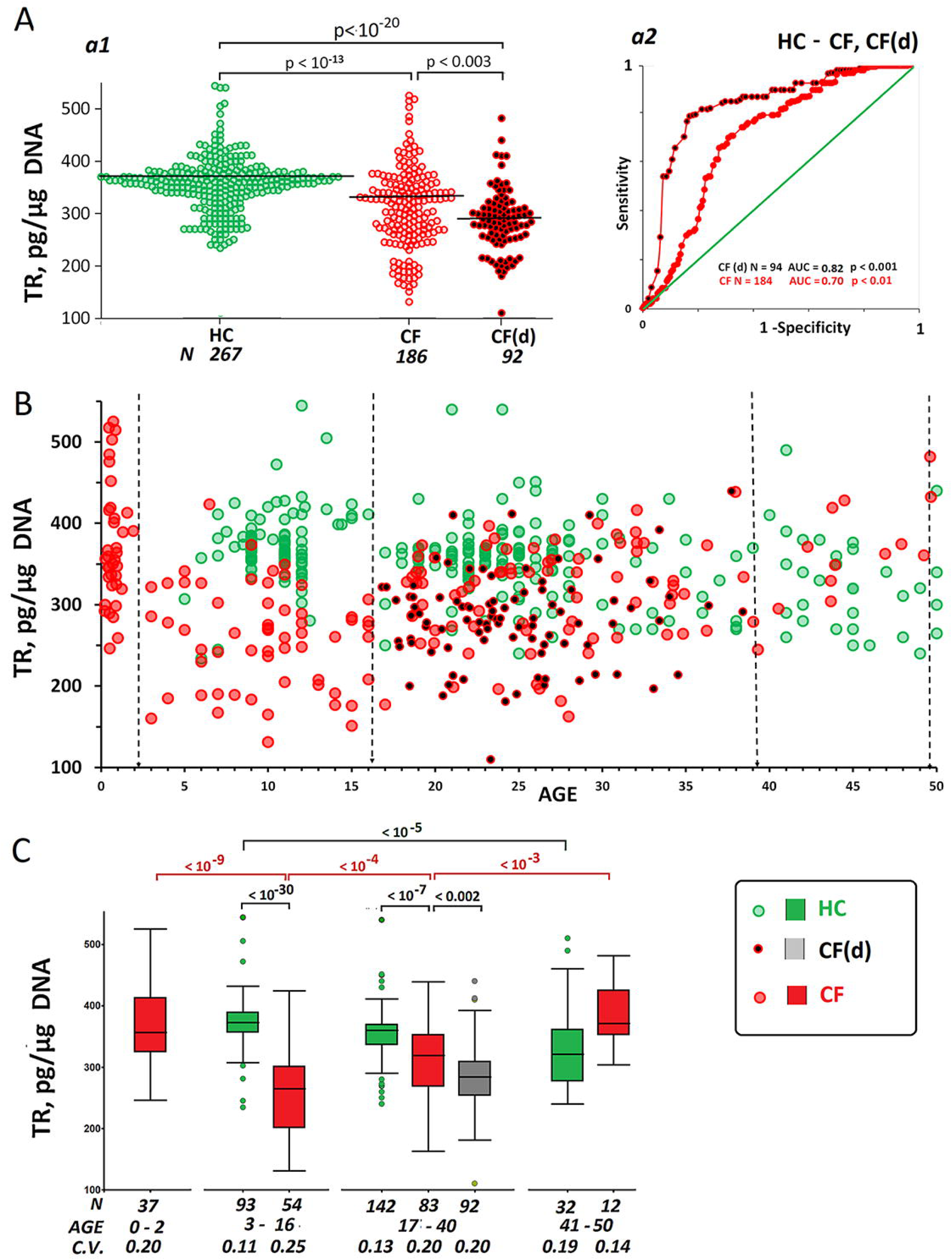
Telomere repeat (TR) abundance in DNA of cystic fibrosis cases and healthy controls. A. (a1) The TR copy numbers in DNA samples from HC, CF and CF(d) groups. A comparison between the groups is reported (U-test). (a2) ROC curves for the groups. B. The dependence of TR copy numbers on subject’s age in HC, CF and CF(d) group. The vertical dotted lines indicate dividing into four age groups. C. The TR CN analysis in different age groups. Box Plot diagrams and group comparisons (U-test) are shown.

**Figure 5.**
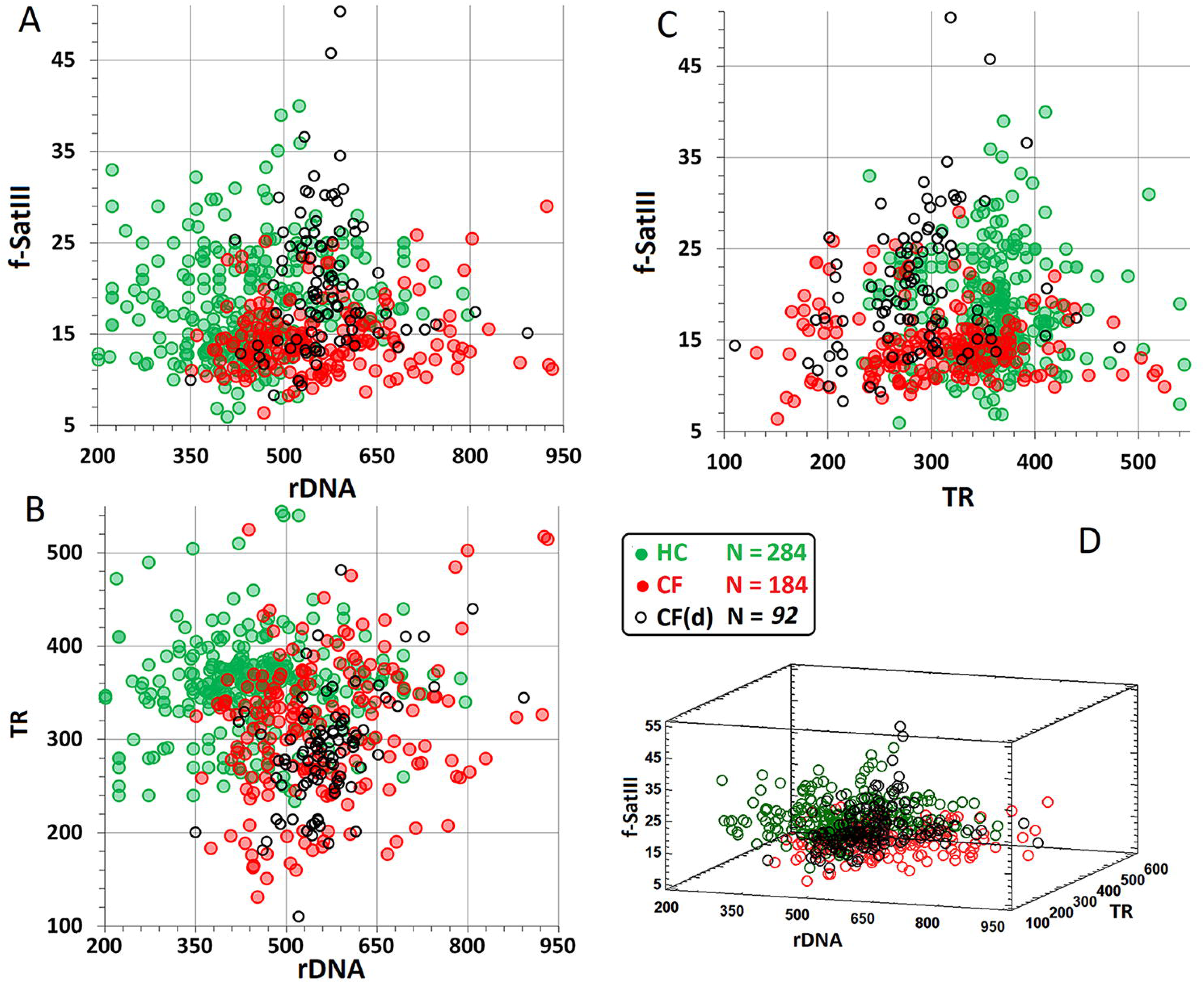
A. A plot of f-SatIII content as function of the content of rDNA. B. A plot of TR content as function of the content of rDNA. C. A plot of f-SatIII content as function of the content of TR. D. A plot of f-SatIII content as function of the content of rDNA (X axis) and TR (Y axis).

70% of the DNA samples from CF(d) group appeared within the following ranges of the three parameters: rDNA CN 500 to 650 copies, f-SatIII 6 to 35 pg/ng DNA, and TR 200 to 400 pg/μg DNA (Fig.5A-D). These ranges comprised merely 35% of DNA samples of the group CF and only 16% of samples of the group HC.

64% of DNA samples of HC group were grouped in the following area: 200 to 500 rDNA CN, 6 to 35 pg/ng DNA f-SatIII, and TR more than 300 pg/μg of DNA. This area comprised 19% of DNA samples of CF group and mere 3% of samples belonging to the most affected CF(d) group.

Thus, DNA samples of affected cases and healthy controls significantly differed by the combinations of the contents of the three repeats. The closest grouping of DNA samples in terms of the contents of the three repeats was characteristic of the CF(d) group with a severe course of the disease and a lethal outcome (Fig.5D).

### 3.5. Analysis of differences in the values of parameters between groups CF and CF(d)

The search for a prognostic marker associated to the disease severity was conducted by comparing parameters (indices) calculated for the patients from CF(d) group aged 17 to 40 years (N=92) and for the age-matched patients from CF(17-40) group (N=83). We analyzed the ratios f-SatIII/rDNA, TR/rDNA and f-SatIII /TR. Indicator rDNA*(f-SatIII/TR) simultaneously takes into account three parameters. The statistics showing the differences in experimental and calculated values of the parameters are listed in Table 3. The greatest differences between the analyzed subgroups CF and CF(d) were observed for the parameters f-SatIII /TR (AUC=0.82) and rDNA*f-SatIII/TR (AUC=0.86, cut-off – 32, specificity – 0.87 and sensitivity – 0.74).

**Table 3.** Comparison of experimentally determined and calculated parameters for CF and CF(d) groups of patients of the same age (17 – 40 y.o.).]

| № | Index | Groups |  | Kolmogorov-Smirnov test |  | ROC- analysis |  | U-test |
| --- | --- | --- | --- | --- | --- | --- | --- | --- |
| | | X1 | X2 | D | $\alpha$ | AUC | p | p |
| 1 | **rDNA** | CF(d) | CF | 0.30 | <0.001 | 0.63 | <0.01 | <0.006 |
| 2 | **f-SatIII** | CF(d) | CF | 0.54 | <10<sup>-10</sup> | 0.79 | <0.001 | <10<sup>-9</sup> |
| 3 | **TR** | CF | CF(d) | 0.37 | <10<sup>-4</sup> | 0.65 | <0.01 | <0.002 |
| 4 | **f-SatIII/rDNA** | CF(d) | CF | 0.37 | <10<sup>-4</sup> | 0.70 | <0.001 | <10<sup>-5</sup> |
| 5 | **TR /rDNA** | CF | CF(d) | 0.47 | <10<sup>-7</sup> | 0.72 | <0.001 | <10<sup>-8</sup> |
| 6 | **f-SatIII/TR** | CF(d) | CF | 0.61 | <10<sup>-13</sup> | **0.82** | <0.001 | <10<sup>-13</sup> |
| 7 | **rDNA\*f-SatIII/TR** | CF(d) | CF | 0.65 | <10<sup>-15</sup> | **0.86** | <0.001 | <10<sup>-15</sup> |

Fig. 6 shows the values of parameters f-SatIII/TR and rDNA*f-SatIII/TR for the three subject groups. The maximum values were found in CF(d). In subgroup CF(3-16), the values were elevated as compared to subgroup CF(0.2-2). The same increase was also found in subgroup HC(17-40) compared to subgroup HC(3-16). In subgroup CF(17-40), index f-SatIII/TR and rDNA*f-SatIII/TR were considerably reduced as compared to subgroup CF (3-16). The analysis of distributions of this parameter corroborated significant differences between groups CF (3-16), CF(d) and the other groups (Fig.6C, data is provided for the index rDNA*f-SatIII/TR).

**Figure 6.**
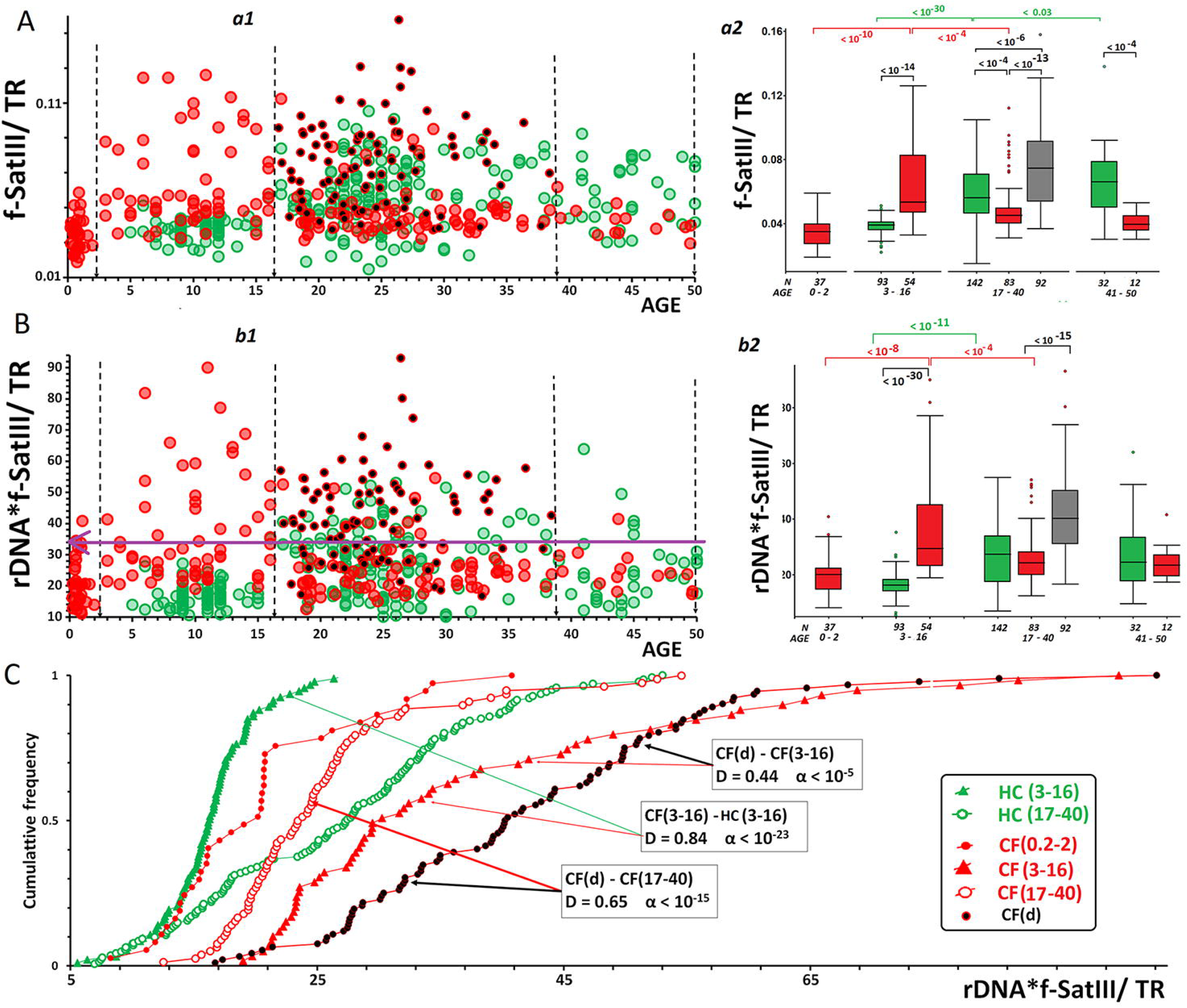
The analysis of calculated parameters f-SatIII/TR (A) and rDNA*f-SatIII/TR (B). (a1, b1) – Relationship between the corresponding parameter and age. (a2, b2) – The analysis of parameter values in different age groups. Box Plot diagrams and group comparisons (U-test) are shown. C. The analysis of rDNA*f-SatIII/TR index distributions in different age groups of CF cases using Kolmogorov-Smirnov test.

### 3.6. Changes in the copy numbers of the three repeats in patients DNA with aging

From the blood leukocytes of 22 patients aged 19 to 40 years, DNA isolation was performed twice with an interval of 5-8 years. DNA was analyzed simultaneously in two samples of blood leukocytes for each patient. Figure 7Aa1-a3 shows the copy numbers of the repeats in two DNA samples of the same patient. The parameter rDNA CN changed in neither patient with the course of time. The parameter f-SatIII increased in 10 patients and decreased in three cases. The parameter TR decreased in 8 cases, while increased in one case only.

**Figure 7.**
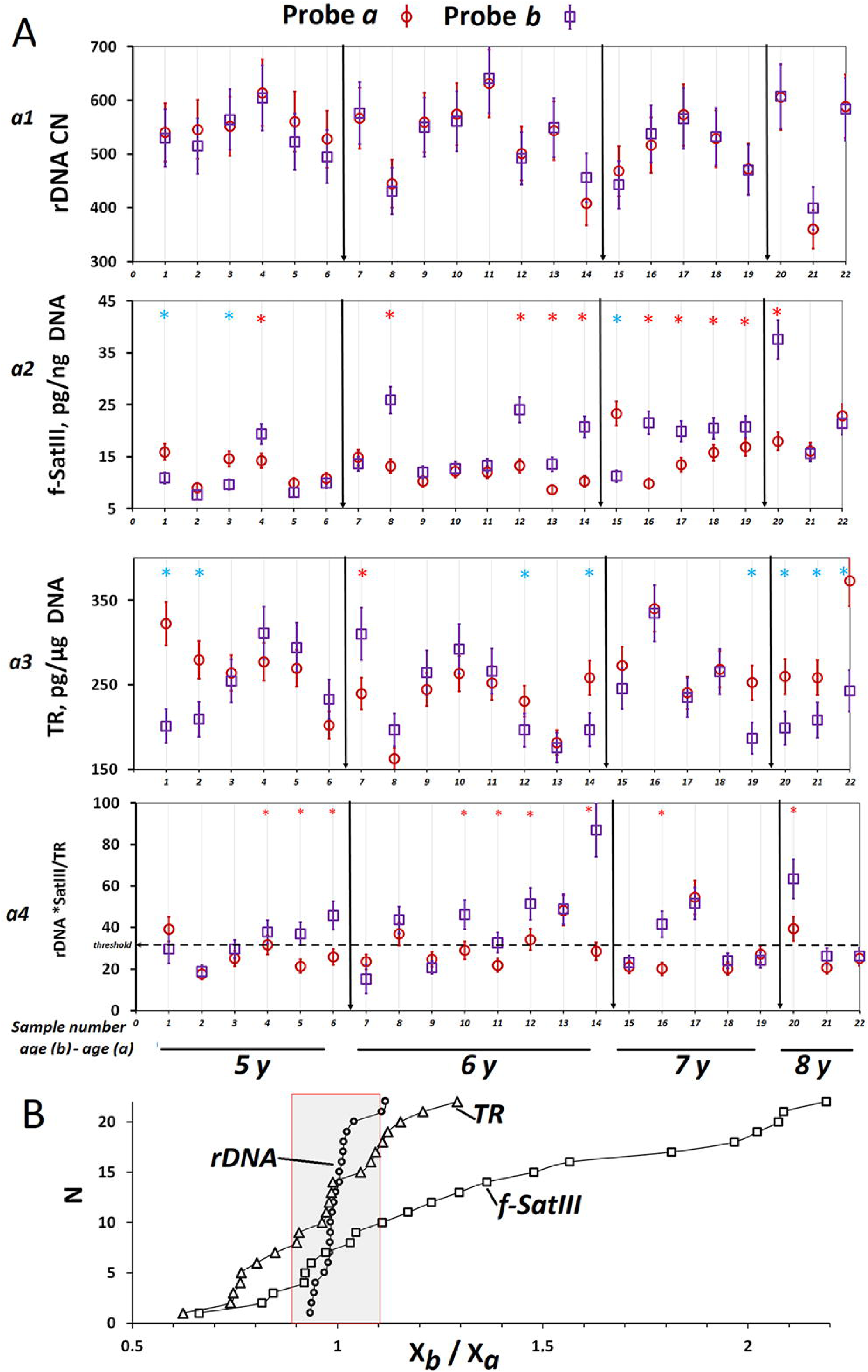
A. Changes of rDNA (a1), f-SatIII (a2), TR (a3) copy numbers and index rDNA*f-SatIII/TR in DNA samples isolated from blood of the same subjects at various time intervals (ΔT, indicated in the caption under X axis). (*)The difference is significant (p< 0.01). B. Distribution of the ratio of the parameter values in the second sample and the first sample obtained from the same subject. The maximum measurement error is indicated (gray box).

Thus, the copy numbers of telomeric and satellite repeats in the DNA of CF patients varied with aging, whereas the ribosomal repeat content remained stable. TR copy numbers tended to decrease, while f-SatIII content tended to increase in DNA with the course of time. Figure 7Aa4 also shows changes in the parameter rDNA*f-SatIII/TR for 22 patients. The dotted line shows the cut-off (32 units) determined via ROC analysis (Fig.6D). The index rDNA*f-SatIII/TR increased in 9 patients over time. In two patients, it reached values of 67 (#20) and 93 (#14), which significantly exceeded the cut-off of 32 units and mignt indicate aggravation in the condition of the patients.

## 4. DISCUSSION

We quantified three types of tandem repeats of the human genome (Fig.1A) in the same DNA samples isolated from the peripheral blood leukocytes of 278 patients with cystic fibrosis and 267 healthy controls. The studied repeats have different biological functions and characteristics indicating their abundance and localization in the nucleus, their CNVs under stress and in the course of human life or during replicative aging of a cell culture.

Our data have shown that rDNA CN is a stable genetic trait that is formed during the formation of a zygote and remains practically unchanged over the lifespan (Fig.7Aa1). The genomes of patients with cystic fibrosis harbored more rDNA copies compared to the controls (Fig.2). Probably, higher rDNA CN values are essential for survival in the presence of the genetic pathology in the genome of the fetus. The prenatal development with the hereditary pathology requires for an elevated protein synthesis (ribosome biogenesis) that only an elevated rDNA CN can provide [62]. In addition, a large enough number of rDNA copies in the nucleolus stabilizes the heterochromatin of the entire nucleus and reduces the risk of rearrangements under stress induced by a pathology or an external impact [31, 32]. The fact that rDNA copy numbers have been significantly less in DNA samples from soon-to-die patients compared to the survived cases is direct evidence of the selection for this trait resulting in better survivability of patients whose genomes initially carried more copies of ribosomal repeats. The variation in the number of ribosomal repeats over the population occurs during zygote formation and is determined by the combinatorics of the 10 pairs of acrocentric chromosomes in meiosis.

Unlike rDNA, the copy numbers of f-SatIII and TR repeats in human cells changes depending on aging and the endogenous and exogenous stress. Fig. 8 represents a scheme illustrating the changes in the contents of the repeats in primary and cultured human cells, which is based on our previous experimental findings [18, 37, 38, 42]. The cell pool is heterogeneous by the abundance of f-SatIII repeats. The fraction of the repeats in bulk DNA isolated from cells depends on the percentage of cells with different f-SatIII content. Most cells of a young body or a cell culture at early passages (type **a**, Fig.8) contain low f-SatIII amounts (Fig.3) and long telomeres (Fig.4). The index **f-SatIII/ TR** for the isolated DNA is minimum (Fig.6). The type **a** cells prevail in leukocyte pools of patients that belong to groups CF(0-2) and HC(3-16).

**Figure 8.**
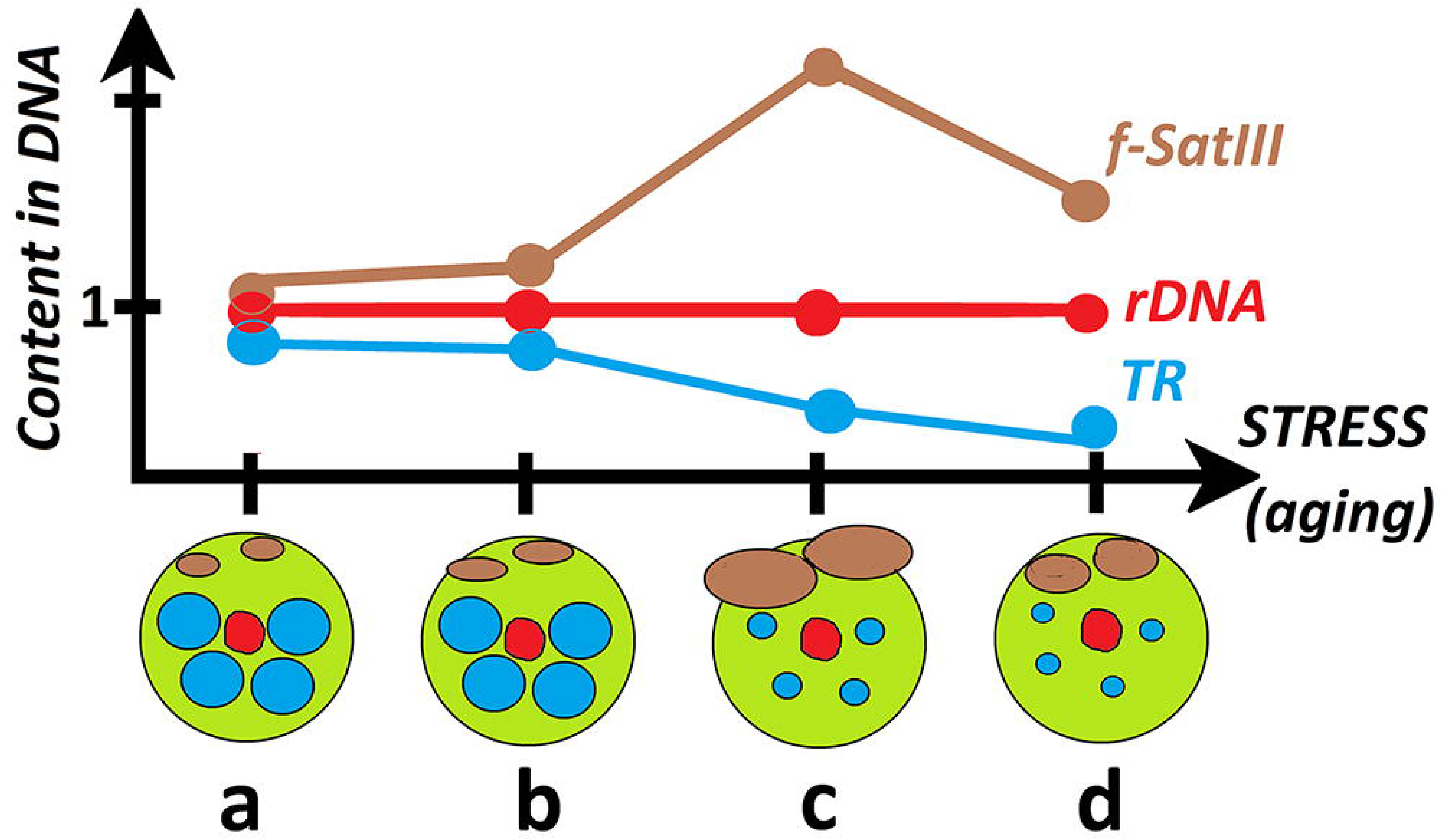
A diagram illustrating changes in the contents of the three repeats in DNA isolated from a cell pool depending on age and/or stress. A, b, c and d are cell types that occur in the cell pool in various proportions depending on age and/or the level of oxidative stress. The diagram is based on the data obtained by the authors earlier.

During normal aging or under stress conditions, a fraction of cells with an elevated f-SatIII content increases in the cell pool [42]. The process that underpins the enlargement of f-SatIII abundance in normal (non-cancer) cells is still not described. For cancer cells, a copy gain scheme has been offered, that includes decondensation of heterochromatin, satellite repeat transcription, reverse transcription of the satellite RNA, and incorporation of DNA/RNA hybrid fragments into the nuclear DNA within pericentromeric loci. The authors suggest that cancer-associated derepression of specific repetitive (satellite) sequences can promote their RNA-driven genomic expansion [41]. The heterochromatin decondensation and transcription activation can promote the telomere shortening. During aging or stress, the cell pool enriches with type **b** and **c** cells (Fig.8).

An increase in f-SatIII content and a decrease in TR are observed in DNA isolated from these cells, while rDNA copy number remains stable. The type **c** cells do not respond to proliferative stimuli, and they are not capable of developing an adaptive response to a stress due to the chromatin rigidity [18]. These cells are eliminated from the pool when stress levels increase. If the fraction of these cells becomes too large and/or the process of their elimination is blocked, the cell population loses its ability to function normally.

In oldest cells or at a high chronic stress level, we observed a decrease in both f-SatIII and TR contents resulting in enrichment of the pool with type **d** cells (low contents of both telomere and satellite repeats in isolated DNA). The decrease in f-SatIII abundance in these cells might be caused by a block of the satellite transcription process. We showed earlier that the level of satellite III transcription depended non-linearly on the dose of the stress factor. Low doses activated the process, whereas high doses blocked it [18].

The cohort of CF patients was heterogeneous in respect of patient age and disease severity. The highest percentage of severe cases should be expected in CF(0-2) and CF(3-16) groups. The groups comprise 97-98% of “severe” *CFTR* mutation cases. The presence of “severe” mutations is known to be associated with an unfavorable prognosis and a shorter life expectancy for patients with cystic fibrosis [63, 64]. Older CF groups comprise fewer “severe” *CFTR* mutation cases. A part of “severe” CF(0-16) patients does not seem to survive and live to adulthood. The copy numbers of f-SatIII and TR depend on two factors: age and stress. The dysregulation of chloride channels results in an elevated level of the oxidative stress [55, 56]. Under the chronic pathology-driven stress the cells age faster. As a result, we observed an increase in f-SatIII/TR index by a factor of 1.5 to 3 in 40% of DNA samples from CF(3-16) group. In controls, this index did not increase before age 17 (Fig.6). Earlier, we observed the same changes in f-SatIII/ TR index during replicative senescence of cultured human skin fibroblasts [42]. Thus, accumulation of type **c** cells occurred in the population of blood leukocytes in 40% of patients over the age of two (Fig.8). The type **c** cells are characterized by a high f-SatIII abundance and short telomeres. Telomere shortening has been reported in blood cells of severe CF patients [65].

The comparison of the three age-matched groups [HC, CF(17-40) and CF(d)] has shown that a feature of survived patients older than 16 years with a relatively favorable course of the disease is a very low content of f-SatIII repeats in DNA compared to the age-matched controls (Fig.3) and deceased patients. We found earlier a similar effect of decreased copy numbers of f-SatIII repeats in DNA of some patients with schizophrenia. Like cystic fibrosis, schizophrenia is associated with chronic oxidative stress [54]. The small fraction of cells with an enormously elevated f-SatIII copy numbers in the population of blood leukocytes of the patients under chronic oxidative stress can be explained by several factors.

Firstly, oxidative stress stimulates the process of elimination of defective cells with an enormously high content of f-SatIII, which are not capable of developing an adaptive response to the stress [18]. Secondly, in the case of schizophrenia, we have shown that some compounds used in therapy are capable of suppressing the transcription processes that potentially result in an increase in the satellite content through the reverse transcription-driven copy gain mechanism described for cancer cells and hypothesized for normal cells. In addition, the compounds had stimulated the process of autophagy in cultured cells, which led to the elimination of cells with a high content of the satellite [37, 38]. Certain drugs used to treat complications of cystic fibrosis could also modulate the processes leading to similar changes in the satellite content of the cells of leukocyte pool. A larger fraction of the cells with elevated f-SatIII abundance in the group of deceased patients CF(d) suggests that too many of these cells are formed due to more active repeat transcription and/or disrupted processes of elimination of these cells from the bloodstream in these patients.

When comparing CF(17-40) and CF(d) groups, we found that an index to detect the differences between the groups in the best way is a parameter equal to the product of the **f-SatIII/TR** index and the content of rDNA (**rDNA** indicator). Large rDNA CNs are associated with a more severe genetic pathology as they can compensate for stress during prenatal and early postnatal life, while low rDNA CNs are eliminated. However, a severe genetic pathology results in a severe disease course in children. The index **f-SatIII/TR** reflects a degree of dysfunction in the cell pool due to the severe disease course induced by a severe mutation or other causes.

ROC-analysis determined the optimum cut-off for the index rDNA*f-SatIII/TR as 32 units (Fig.6). Within CF(d) group, the index exceeded this cut-off theshold in 74% of DNA samples. In CF(3-16) subgroup, a fraction of such DNA samples was 51%, while in CF (0-2) and CF(17-50) subgroups, such samples formed merely 9%. The indices rDNA*f-SatIII/TR and f-SatIII/TR might be used as a prognostic marker of the expected lifespan of a cystic fibrosis patient.

The processes that underpin the CNVs of f-SatIII repeats are of great importance for the essential body functions during aging and/or in pathology and require further research. The **c** type cells (Fig.8) are a ballast for the cell population because they do not respond to various stimuli, including those inducing proliferation or a normal adaptive response. The inability of the cells to respond to stimuli requiring alteration of the genome expression profile is associated with inability to change the structure of chromatin in response to the stimuli [18]. Perhaps, a therapy aimed at reducing the fraction of f-SatIII in the genomes of the cases would help improve the condition of the patients with the severe course of the disease.

## 5. CONCLUSION

Cystic fibrosis (CF) was associated with higher rDNA abundance and altered f-SatIII and TR contents in the DNA of cases compared to the controls. The severe course of the disease was characterized by high f-SatIII contents and shortened telomeres. The mild course of CF was associated with low contents of f-SatIII and normal or slightly reduced telomere length. The index **rDNA*(f-SatIII/TR)** might be a predictor of the patient’s life expectancy.

## Data Availability

All data produced in the present study are available upon reasonable request to the authors

